# Evaluating strategies for spatial allocation of vaccines based on risk and centrality

**DOI:** 10.1101/2021.09.07.21263209

**Authors:** Benjamin J Singer, Robin N Thompson, Michael B Bonsall

**Affiliations:** Department of Zoology, University of Oxford; Mathematics Institute, University of Warwick; Zeeman Institute for Systems Biology and Infectious Disease Epidemiology Research, University of Warwick

**Keywords:** vaccine strategy, network centrality, epidemic modelling, metapopulation modelling

## Abstract

When vaccinating a large population in response to an invading pathogen, it is often necessary to prioritise some individuals to be vaccinated first. One way to do this is to choose individuals to vaccinate based on their location. Methods for this prioritisation include strategies which target those regions most at risk of importing the pathogen, and strategies which target regions with high centrality on the travel network. We use a simple infectious disease epidemic model to compare a risk-targeting strategy to two different centrality-targeting strategies based on betweenness centrality and random walk percolation centrality, respectively. We find that the relative effectiveness of these strategies in reducing the total number of infections varies with the basic reproduction number of the pathogen, travel rates, structure of the travel network, and vaccine availability. We conclude that, when a pathogen has high spreading capacity, or when vaccine availability is limited, centrality-targeting strategies should be considered as an alternative to the more commonly used risk-targeting strategies.

## 1 Introduction

Administrators of vaccination programmes often find that there are fewer doses of vaccines available (or capacity to administer these doses) than there are susceptible individuals willing to receive them. In this situation, some form of prioritisation is necessary, in which certain individuals are chosen from the susceptible population to be vaccinated, or to receive vaccines earlier than others. Often, vaccines are offered to individuals in certain demographics who are particularly at risk of catching, spreading, or experiencing severe outcomes of a disease. This kind of demographic targeting is used by the United Kingdom’s National Health Service to distribute influenza vaccines—the NHS offers free vaccines to individuals over 65, or with conditions such as asthma and diabetes, who are at risk of severe influenza [1]. Targeting demographics at high risk of severe disease has also been a key strategy in COVID-19 vaccination programmes, with elderly people often being prioritised [2, 3, 4].

An alternative way of selecting which individuals receive vaccinations is by virtue of their location in geographic space. Spatial position can be used as an alternative to demographic membership as a predictive variable for risk of infection [5], but often spatial strategies are used when cases of a disease occur only within a certain region, and public health decision-makers seek to contain the pathogen locally and prevent it from spreading to other regions [6, 7, 8]. Spatial control can be particularly important when responding to pandemic risks at the point when containment is still possible [9]. While we focus on vaccination to provide a concrete example of a spatial control tool, the structure of our model is general and can represent any prophylactic control strategy that prevents infections (e.g. administering infection-preventing drugs, or culling of animals and inducing resistance in plants in the case of agricultural diseases).

Within spatial strategies, there are many ways in which certain regions can be prioritised for vaccine distribution. Among alternatives, priority could be given to regions close to an existing epidemic, with high population density, or with many travel connections. In practice, many modelling studies that consider spatial vaccine allocation use a risk-targeting approach, in which those regions predicted to be most at risk of importation of a pathogen are prioritised for distribution of the vaccine [5, 8, 10]. There is, however, no proof that this leads to the maximum reduction in infections, and authors often mention possible further work examining “downstream effects” [10] or “spreading capacity” [11] of certain regions, or the “impact of vaccination on … spatial expansion”’ [5].

Here we define a vaccine strategy as a method for ordering all individuals susceptible to an infection from highest priority to be vaccinated, to lowest priority. We define a vaccine allocation as a division of those individuals into two sets—those in one set receive a dose of vaccine, and those in the other do not. We define risk-targeting as any vaccine strategy in which a region is prioritised for receipt of vaccine doses based solely on the risk that a large outbreak of the relevant pathogen will occur in that region. The method for determining this risk can range from simple heuristics to sophisticated epidemiological forecasts [8]. One particularly widely discussed form of risk-targeting is spatial ring vaccination (as opposed to ring vaccination of contacts), in which individuals are prioritised for vaccination based on their proximity to an ongoing outbreak. This kind of strategy has been discussed in settings from foot-and-mouth disease in the United Kingdom to Ebola in the Democratic Republic of the Congo [6, 7].

An alternative to risk-targeting in strategies for vaccine distribution is centrality-targeting [12, 13, 14]. Centrality is a term for any measure of the importance of a node in a network, and it can be used as a proxy for the spreading capacity of a given region. We discuss specific measures of centrality in more detail below. In centrality-targeting strategies, vaccines are preferentially allocated to regions with high centrality in a travel network. Thus, they may act to impede the circulation of a pathogen between regions, rather than directly protecting those regions most likely to experience outbreaks themselves.

There is little existing work directly comparing risk-targeting of vaccines to centrality-targeting. A study by Piraveenan et al. (2013) compares targeting of prophylactic control in a disease transmission network according to three different measures: hop distance (a simple kind of risk-targeting), percolation centrality, and betweenness centrality [12]. Hop distance is the length of the shortest path through the network from a susceptible node to the closest infected node, and can be treated as a proxy for risk. Betweenness centrality is a very commonly used centrality measure based on a node’s connectivity. Percolation centrality is a measure of node centrality of the authors’ own devising, which takes into account a node’s connectivity as well as the current state of infection in the network. The authors find that, in a scale-free and a random network, which of the three strategies best reduces infections depends on parameters reflecting how widespread the pathogen is, and how many nodes can be immunised. In general, when the pathogen is scarce in the network, or when many nodes can be immunised, risk-targeting (that is, priority according to hop distance), is most effective at reducing infection. In the opposite scenario, when the pathogen is widespread or when few nodes can be immunised, betweenness-targeting is most effective. In the intermediate regime, targeting according to percolation centrality led to the largest expected reduction in infections.

Generalising from this result, we conjecture that risk-targeting may be less effective than centrality-targeting when resources available for control of a pathogen are very limited (e.g. few doses of a vaccine), or when the pathogen concerned has a high spreading potential (either by having a high *R*_0_ or by travelling widely across space). We argue that Piraveenan et al.’s findings are intuitive—for instance, when many resources are available for control, a pathogen can be fully contained by ring vaccination (a form of risk-targeting), while the only chance to contain a pathogen when few resources are available is by targeting travel hubs (a form of centrality targeting). This intuitive understanding of the scenario leads to the following generalised hypothesis:

- when resources for control are widely available, and the potential for widespread pathogen transmission is limited, targeting regions at high risk is more effective than targeting regions with high centrality.
- when resources are limited and the pathogen has the potential to spread widely, targeting regions with high centrality is more effective than targeting regions at high risk.

To investigate this hypothesis, we examine whether the risk-targeting strategy is the most effective strategy to reduce the overall number of infections in a range of different epidemiological networks. We compare this strategy to two variations on a centrality-targeting strategy that prioritises regions for vaccination according to their centrality on the human travel network. The variations come about due to alternative centrality measures.

## 2 Methods

### 2.1 Disease model

In this study we make use of a simplified stochastic SIR metapopulation model similar to others used in spatial analyses of infectious disease epidemics and their control [5]. Each node *i* on the travel network is associated with a region, which has its own population size *N*_*i*_ and within-region basic reproduction number *R*_0,*i*_. We can represent the travel network with matrix ***λ***, in which the element *λ*_*ij*_ is the expected number of individuals travelling from region *i* to region *j* per unit time. Like all rates referred to in this section, we understand this as a stochastic rate, where the actual number of individuals travelling over a time period Δ*t* is given by a Poisson distribution with rate parameter *λ*_*ij*_Δ*t*.

With this information we can calculate the expected number of infected individuals travelling from a region *i* that experiences an epidemic, to any other region connected to it on the travel network. First we calculate the total number of infected individuals over the course of the epidemic in region *i, R*_*i*_(∞), using the widely used SIR final size equation [15, 16, 17, 18, 19]:

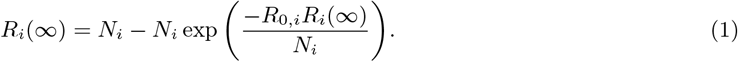

Each infected individual can either recover while still in region *i*, or travel to a neighbouring region before recovering. The probability *P*_*ij*_ that an individual travels to a particular neighbouring region *j* while infected is therefore

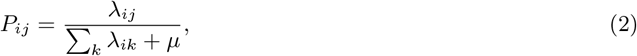

where *µ* is the recovery rate, and Σ_*k*_ *λ*_*ik*_ is the total rate of travel out of region *i*. This means that the expected total number *I*_*ij*_ of infected individuals who arrive in region *j* from region *i* over the course of the epidemic in *i* is

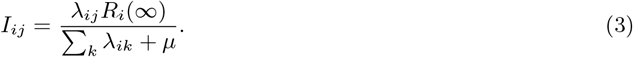

When *µ* ≫ Σ_*k*_ *λ*_*ik*_, meaning that individuals are much more likely to recover than travel while ill, equation 3 can be approximated as:

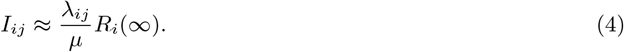

This approximation is motivated in part by the fact that, for the metapopulation model to be appropriate, the time-scale of the dynamics within each sub-population must be much faster than the time-scale of mixing between the populations. If the two time scales were similar, it would not be appropriate to represent the sub-populations as separate systems. We also make use of this approximation to assume that the population size *N*_*i*_ of each region does not change over the course of the outbreak.

To avoid highly intensive computations, let us consider a simplified disease model with a square epidemic curve—meaning that, in our model, there are a constant number of infected individuals during an epidemic, and no infected individuals otherwise. We assume that the duration of such an epidemic is exponentially distributed, but that the number of infected individuals is deterministic. Since the size of the epidemic does not vary from its start to its end, this means that we can shift from treating the states of individuals, to treating the states of whole regions. We can treat each region as belonging to a state *S*, meaning that no epidemic has occurred; *I*, meaning that an epidemic is ongoing; or *R*, meaning that an epidemic has occurred and the region is protected from further outbreaks due to herd immunity.

The expected total number of infected individuals who arrive in region *j* from a region *i*, which experiences a square epidemic curve with height *A* and mean duration *D*, is given by *λ*_*ij*_*AD*. This quantity determines the probability for the pathogen to spread between regions, so we seek to match its value in our simplified model to the corresponding value in equation 4. To achieve this, we choose *A* = *R*_*i*_(∞)*/µ* and *D* = 1. This choice ensures that one unit of time is equal to the expected duration of a single epidemic, thus effectively rescaling time to match the simplified pathogen dynamics.

In a stochastic SIR model, the probability for a single infected individual to cause an epidemic when introduced into a large, entirely susceptible population is 1 − 1*/R*_0_, a result cited widely in the epidemiology literature [16, 20, 21, 22, 23, 24]. So if the rate of arrival of infected individuals in region *j* from region *i* is *λ*_*ij*_ *R*_*i*_(∞)*/µ*, and each arrival has a probability of causing an epidemic given by 1 − 1*/R*_0_, then the rate at which the epidemic in region *i* seeds an epidemic in region *j* is

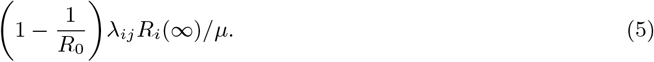

This completes the structure of our model: regions are assigned states *S, I*, and *R*; regions in state *I* export the epidemic stochastically at the above rate to neighbouring regions in state *S*; and regions in state *I* recover stochastically into state *R* at rate 1, after which they cannot be reinfected. We assume that the time between events is exponentially distributed, and use Gillespie’s direct stochastic simulation method to generate our results [25].

### 2.2 Centrality measures

The alternatives to risk-targeting that we investigate are centrality-targeting strategies. Centrality is a measure of the importance of a node in a network. Targeting nodes for intervention based on their centrality may help prevent wide circulation of a pathogen in a transmission network. There are many different ways of measuring centrality, one of the most widespread being betweenness centrality.

The betweenness centrality of a node *v* is the proportion of shortest paths from a source node *s* to a target node *r* that pass through *v*, averaged over all sources and targets *s* and *r* [**?, ?**]. The betweenness *b*_*v*_ of node *v* in a network *G* of size *n* is expressed as

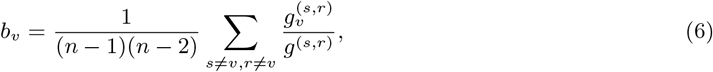

where *g*^(*s,r*)^ is the number of geodesics (shortest paths) from node *s* to node *r*, and 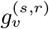 is the number of such paths that pass through node *v*. This definition ensures that the betweenness of a given node is always between 0 and 1.

Betweenness centrality is discussed widely in studies of spreading processes on networks [26, 27, 28, 29]. For this reason we include it in our analysis. However, there is good reason to believe that betweenness centrality is not particularly well-suited to finding important nodes in a disease transmission network [30]. First, it only counts geodesic paths—but infections transmitting between regions have no sense of direction, and so cannot be assumed to follow the shortest path from their source to their destination. Second, when responding to an invading pathogen, we often know the possible sources of an infection and are seeking to disrupt transmission from these sources—whereas the betweenness centrality calculation averages across all possible sources. We can resolve the first issue with random walk betweenness centrality, and the second issue with percolation centrality—combining the two results in our random walk percolation centrality measure.

Random walk betweenness centrality is defined similarly to betweenness centrality, but expands the paths counted, from only shortest paths from node *s* to node *r* that pass through node *v*, to all random walks that pass through *v* from *s* to *r* [31, 32]. Since a random walk can have an arbitrarily high number of steps, we allow steps passing along an edge in opposite directions to cancel out, so that we ignore random walks that go back and forth through a given node many times without progressing to another part of the network. We can then define the random walk betweenness *rwb*_*v*_ of a node *v* in terms of the net number of times a random walk from *s* to *r* is expected to pass through 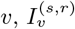.

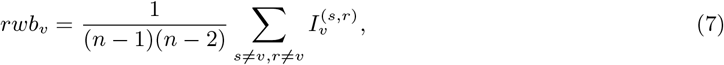

The method for calculating 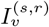 is given in Newman 2005 [31].

Percolation centrality introduces a weighting to each element of the sums in equations 6 and 7 depending on the infection state of the source node *s* [12]. The percolation centrality *p*_*v*_ (*t*) of node *v* at time *t* is

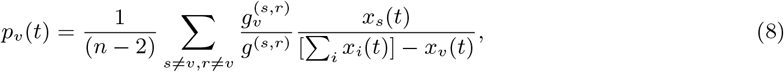

where *x*_*i*_(*t*) is a scalar quantity representing the infection state of node *i* at time *t*. In our analysis we assume that a given region is either infected (state *I*) or uninfected (state *S* or *R*), restricting the values of *x*_*i*_(*t*) to 0 if region *i* is in state *S* or state *R*, and 1 if region *i* is in state *I*. In this binary case, equation 8 can be interpreted as equivalent to equation 6, with the source node *s* restricted to the class of infected nodes, and the quantity normalised appropriately. This successfully captures the importance of paths starting from an infected node in a disease transmission network.

Combining the modifications to betweenness centrality introduced in equations 7 and 8 gives us random walk percolation centrality. The random walk percolation centrality *rwp*_*v*_ (*t*) of a node *v* at time *t* is

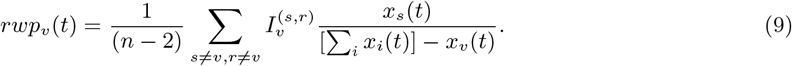

Note that this equation is to equation 7 as equation 8 is to equation 6.

To apply this measure to human mobility networks we need to capture the fact that the edges of these networks are weighted by the amount of travel that occurs along them, and can be directed if travel is not symmetric between regions. In the supplementary information, we present the novel mathematics of random walk percolation centrality in a weighted directed network. This results in the measure of random walk percolation centrality (RWPC) used in this study.

### 2.3 Network specifications

We tested vaccination targeting strategies for simulated epidemics on metapopulations with a variety of transport networks. We chose two network structures, two artificial and two empirical. Our artificial networks are based on a “joined grids” structure, in which the regions are split into two identical subsets, each of which is arranged in a grid with unit distance between orthogonally adjacent regions (see figure 1). Travel occurs at the highest rate between regions (represented by nodes) which are adjacent on the grid, and at a rate *λ*_*ij*_ determined by distance between more distant regions. In particular:

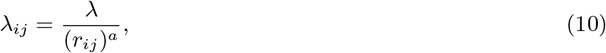

**Figure 1:**
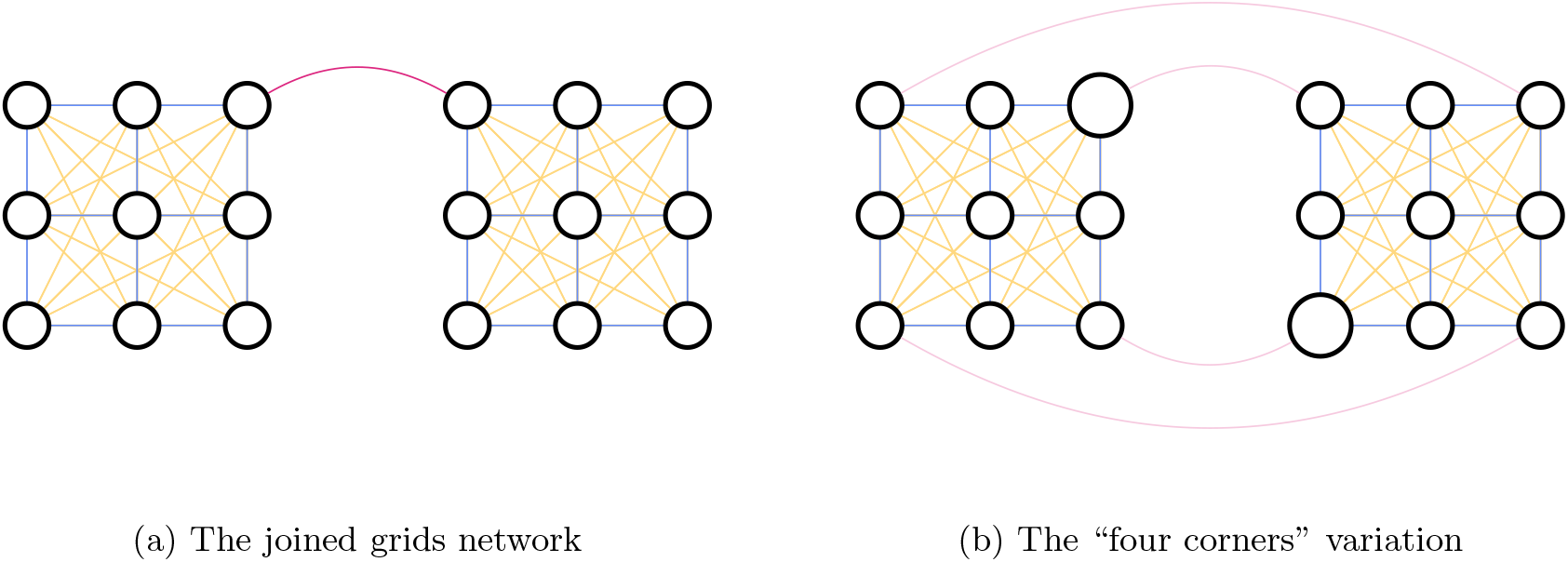
The “joined grids” arrangements of regions (circles), in which the movement of the population happens between adjacent regions in each grid (blue), on longer-range travel within a grid (yellow), or along inter-grid travel routes (red). The area of the circles scales with the population of the corresponding region.

where *λ* is the travel rate between adjacent regions, *r*_*ij*_ ≥ 1 is the distance between region *i* and region *j*, and *a* is a parameter determining the amount of long-distance travel—the higher the value of *a*, the less long-distance travel. This is a kind of gravity model, which is one of the standard models of human movement [35, 36, 37, 38]. In the basic version of this network, illustrated in 1a, there is a single travel route between the two grids, which runs from a region at the corner of one grid to a region at the corner of the another. We set the travel rate along this route at *λ*_*join*_ = *λ* by default. With this model we can represent outbreaks occurring at three different scales—within regions (represented by circles in figure 1), within grids (each grid is made up of nine regions), and across the whole network (the whole network is made up of 18 regions on two grids). We assume that each region has a fixed population of 200,000.

To explore further the role of population size and alternate routes of transmission in an artificial network, we created a variation on the basic joined grids arrangement, which we call the “four corners” network. In this network, the travel within each grid is given by equation 10, but there are four routes between the two networks, joining each corner of each grid to a corner of the the other grid, as illustrated in figure 1b. The inter-grid routes each have travel rate *λ*_*join*_ = *λ/*4. Each region has a fixed population of 200,000, except for one region on the corner on each grid, which has a population of 400,000, to represent high-population travel hubs. These two regions are not connected by any direct travel route. To account for the greater travel to more populous regions predicted by the gravity model of human movement, we now write

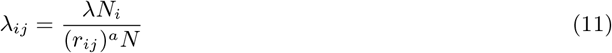

for the within-grid travel rates, where *N* = 200, 000.

To extend our theoretical results to real-world networks, we make of two freely available data sets concerning human movement patterns. The first is a subset of the US domestic air traffic network, sourced from the Bureau of Transport Statistics [39]. We used as nodes in our network only the 20 airports with the highest total sum of passengers arriving and passengers departing, and defined the weight of a directed edge between two of these nodes *i* and *j* in proportion to the total number of passengers travelling from airport *i* to airport *j* in 2019. To assign a population size to each node, we used the population size of the urban area in which the airport is situated [40]. There are two airports in the data set situated in Chicago. In this case, we split the population equally between the two airports, assigning half the total population of the Chicago urban area to each node.

The edge weight *λ*_*ij*_ (i.e. the travel rate) from node *i* to node *j* in this network is therefore given by:

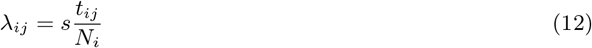

where *t*_*ij*_ is the total number of passengers travelling from airport *i* to airport *j* in 2019, *N*_*i*_ *>* 0 is the population size of node *i*, and *s* is an overall scaling factor that we allow to vary.

The second empirical network we use is sourced from the 2011 UK census [41]. We use the census to estimate the amount of travel between all cities and towns with populations over 100,000 in an area of Northwest England including Greater Manchester and West and South Yorkshire (see figure 2b). The census includes data for the location of the residence and place of the work of all respondents, which we use to estimate the number of people *c*_*ij*_ who commute between any given pair of cities *i* and *j*. We use this to model the travel rate according to the equation:

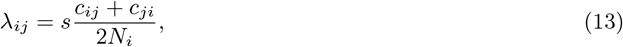

**Figure 2:**
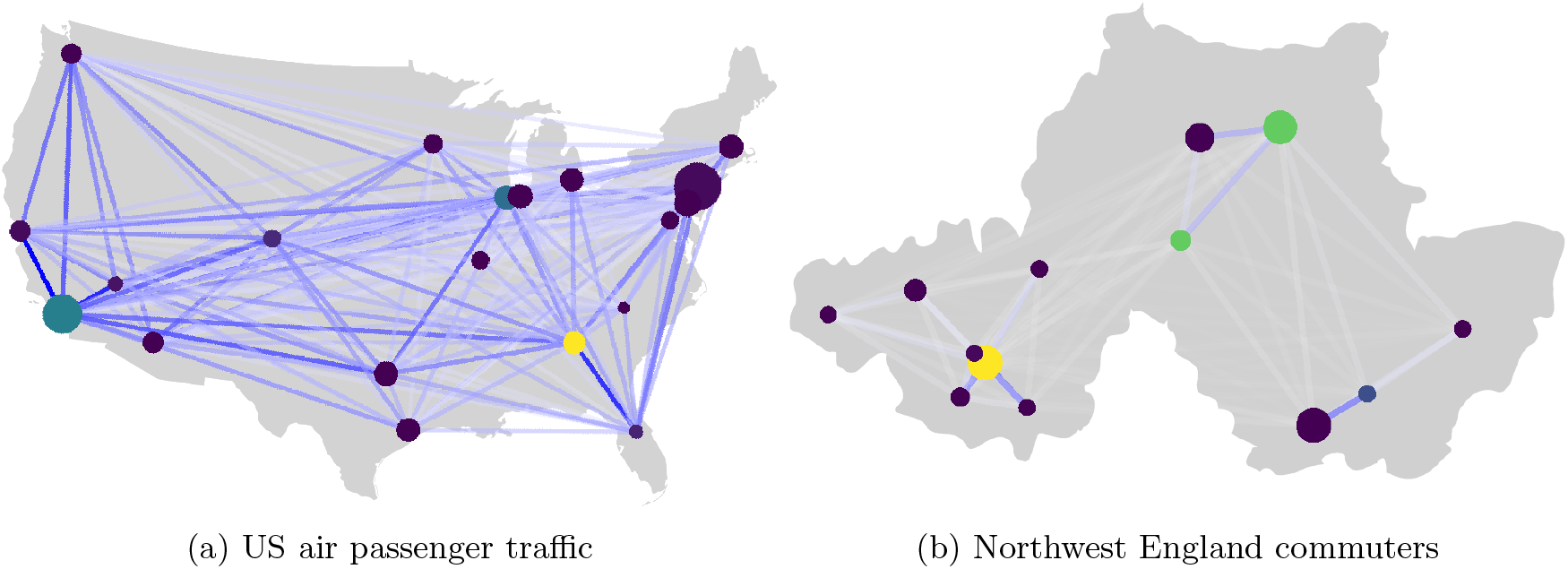
Two travel networks based on empirical data. The colour and opacity of the edges scales with their weight, normalised separately in each network relative to the highest weighted edge. The area of each node scales with the population size it represents, and the colour indicates its betweenness centrality. Chicago has two airports in the US air passenger traffic data set, so these nodes are represented with some jitter. [33, 34]

where *N*_*i*_ *>* 0 is the population size of city *i*, and *s* is an overall scaling factor which we allow to vary. We make the travel rate symmetric because a commuter travelling between cities can transmit a pathogen either from the source city to the destination city (on the outbound journey) or from the destination city to the source city (on the inbound journey) [42]. The key assumption of our models of empirical travel data is that total travel is proportional either to air travel or to commuter travel, respectively.

## 3 Vaccination Strategies

To test different vaccination strategies, we followed three alternative methods of prioritising regions for vaccination—risk, betweenness centrality, and random walk percolation centrality (RWPC). We assumed that a pathogen would emerge in a single region at first, putting that region in state *I* and triggering a vaccination campaign across the network. We performed separate simulations for each possible region of origination of the outbreak, and averaged the results.

We modelled the results of these prioritisations with a range of total available vaccine doses, considering all regions in state *S* as possible targets for vaccination. We assumed a single-dose vaccine with 100% efficacy. Given a certain number of doses *n*_*d*_, we assigned enough doses to the highest priority region to achieve herd immunity in that region, then did the same for the second highest priority region, and so on, until we reached a region whose herd immunity threshold was higher than the number of doses remaining, in which case we assigned all of the doses remaining to that region and none to those with lower priority. We assume that the vaccine distribution in a region is always completed before an epidemic occurs in that region.

For the risk-targeting strategy, we performed 10,000 simulations of the outbreak from the initial state, and prioritised regions from most at risk to least at risk. In this study we treated the risk to region *i* as equivalent to the probability that region *i* would become infected. In principle, the risk is the expected number of infected individuals, i.e the probability of the region becoming infected multiplied by *R*(∞). However, both *R*(∞) and the number of vaccines required to achieve herd immunity scale linearly with the size of the susceptible population. This means that the risk averted per dose of vaccine is maximised by prioritising regions based on the probability of an outbreak in that region, from highest probability to lowest. For the RWPC strategy, we calculated the RWPC for each region according to the initial state of each region, and prioritised regions from highest RWPC to lowest. For the betweenness centrality strategy, we simply prioritised from highest betweenness centrality to lowest betweenness centrality. For both centrality measures we found that weighting the edges according to the rate of travel per individual resulted in closer to optimal targeting than weighting them according to the total flux of travelling individuals.

## 4 Results

The results of vaccine distribution on our artificial networks using all three different strategies is shown in figure 3. In each panel, the y-axis shows the expected number of infected individuals resulting from an epidemic that starts with a random region in state *I* and the remaining regions in state *S*. The lower the number of infected individuals, the more effective the vaccination programme. Note that the scale varies from panel to panel, to allow the variation in effectiveness to be seen clearly in each case. The x-axis in each panel shows the number of vaccines administered, *n*_*d*_, representing the resources available for control. The three different coloured lines show the expected number of infected individuals when vaccines are distributed according to the risk-targeting strategy (blue), the betweenness centrality strategy (red), or the RWPC strategy (yellow). The panels are arranged left-to-right with increasing values of within-region basic reproduction number *R*_0_, and top-to-bottom with increasing values of adjacent-region travel rate *λ*. The range of parameters for which simulation results are shown is chosen to illustrate the region around a change in the relative effectiveness of different vaccine strategies, if such a change occurs.

**Figure 3:**
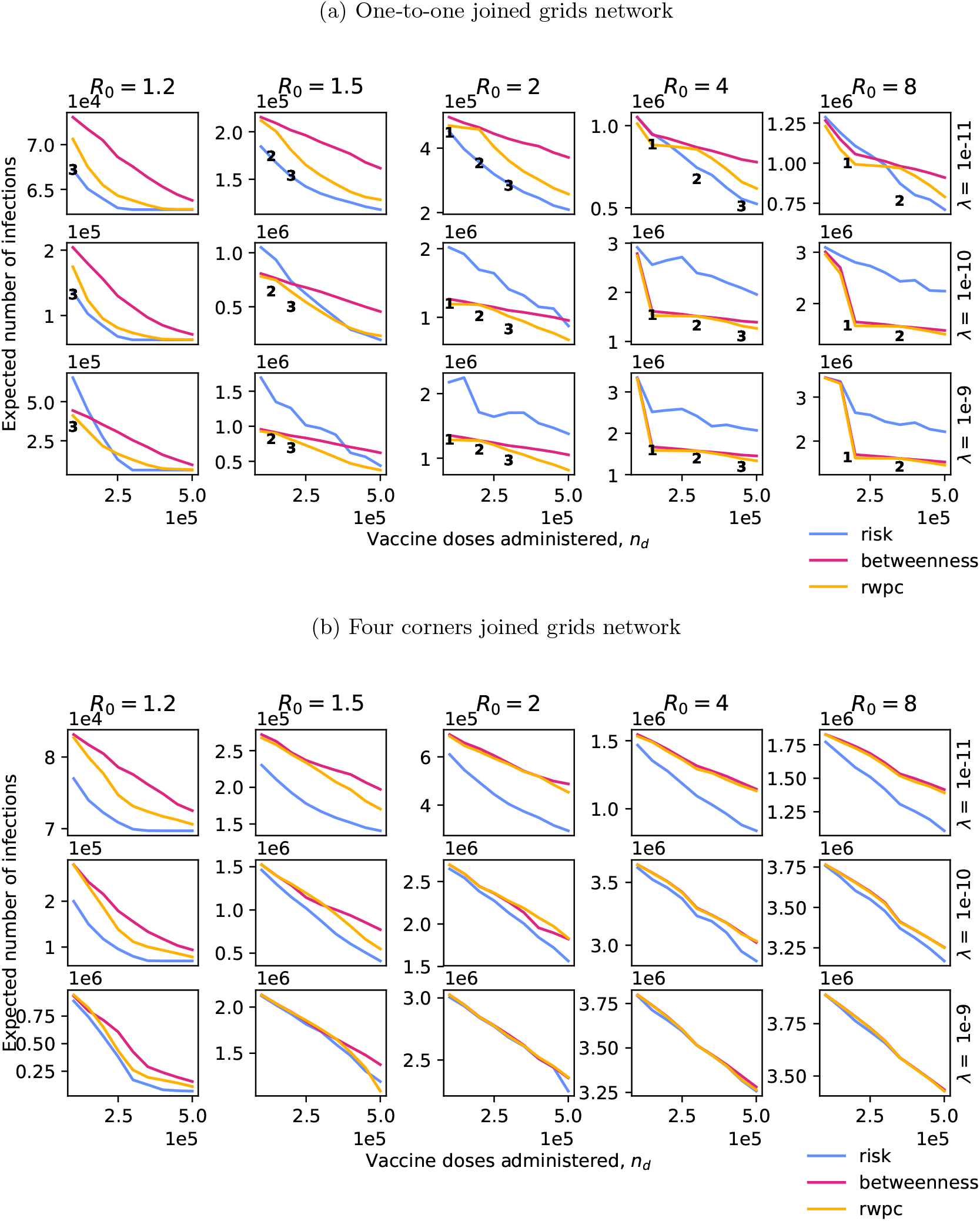
Plots of the expected total number of infections due to a disease outbreak against the number of vaccine doses administered according to one of three region prioritisation policies, for five different values of *R*_0_ and three different overall travel rates, in two variations on the ‘joined grids’ network. The long-distance travel parameter *a* in equation 10 is set to 2 in all cases. In all cases we set *µ* = 0.5. For the one-to-one variation, it is possible to calculate the optimal vaccine allocation for some parameter values. The resultant expected numbers of infections are shown by the positions of the black numbers, which indicate the number of regions vaccinated to herd-immunity at that value of *n*_*d*_.

In figure 3a, we see that, for the one-to-one joined grids network, risk-targeting is the best strategy under some parameter combinations and not under others. When both *R*_0_ and *λ* are small, the risk-targeting strategy is always the most effective of the three considered, for any number of available vaccines. For some combinations of *R*_0_ and *λ*, such as 8 and 2e-6, respectively, the risk-targeting strategy is only the most effective when there are many vaccine doses available. Finally, when both *R*_0_ and *λ* are large, the centrality-targeting strategies are more effective than the risk-targeting strategy at all values of *n*_*d*_. The RWPC strategy is always more effective than the betweenness centrality strategy, with the difference between the two being largest when *R*_0_ and *λ* are small and *n*_*d*_ is large. The large dips in expected infections seen in the lower right panels of the figure correspond to the point at which sufficient vaccines become available to achieve herd immunity in the patch that joins the two grids. This happens outside of the range of doses considered when *R*_0_ ≤ 2, and does not have as dramatic an effect when *λ* = 10^*−*11^. In the supplementary information we show that increasing *a* (and thus decreasing long-distance travel) makes risk-targeting more effective, but does not change the general pattern of the results.

In the joined grids network, where each region has the same population size, it is sometimes computationally feasible to determine the optimal allocation of vaccines using a brute force approach. That is, the approach in which we simulate pathogen spread under all possible vaccine allocation prioritisations, and select the prioritisation that leads to the smallest expected number of infections. For this approach, we constrain the number of doses to be equal to a small integer multiple of the herd immunity threshold for a single population, so that the number of possible allocations is not excessively high. In this case, we assume that no less than the number of doses required for herd immunity are ever allocated to a region, if any doses are allocated there at all. The resultant expected number of infections for each of the optimal allocations is indicated with bold integers in figure 1a, marked by the number of regions vaccinated. The spacing between these numbers scales with the herd immunity threshold, as determined by *R*_0_. In general, the best strategy out of those considered for any combination of parameters comes close to optimality in effectiveness, with risk-targeting at low travel rates being indistinguishable from the fully optimal strategy.

In figure 3b, we see that these dynamics can be significantly altered by changing the network structure. When we add multiple routes between the two grids, and increase the population size of two patches, the risk-targeting strategy becomes the most effective strategy for almost all parameter combinations. For very high spread rates the pathogen infects all those not protected by the vaccine or by regional herd immunity, and so the expected number of infections becomes the same linear function of the number of doses for all vaccine strategies.

In figure 4, we show the results of our various vaccine strategies in empirically derived networks. The structure of the figure is identical to figure 3, but with the scale of travel indicated by the *s* parameter rather than *λ* (see equations 12 and 13 for the role of *s* in figures 4b and 4a, respectively).

**Figure 4:**
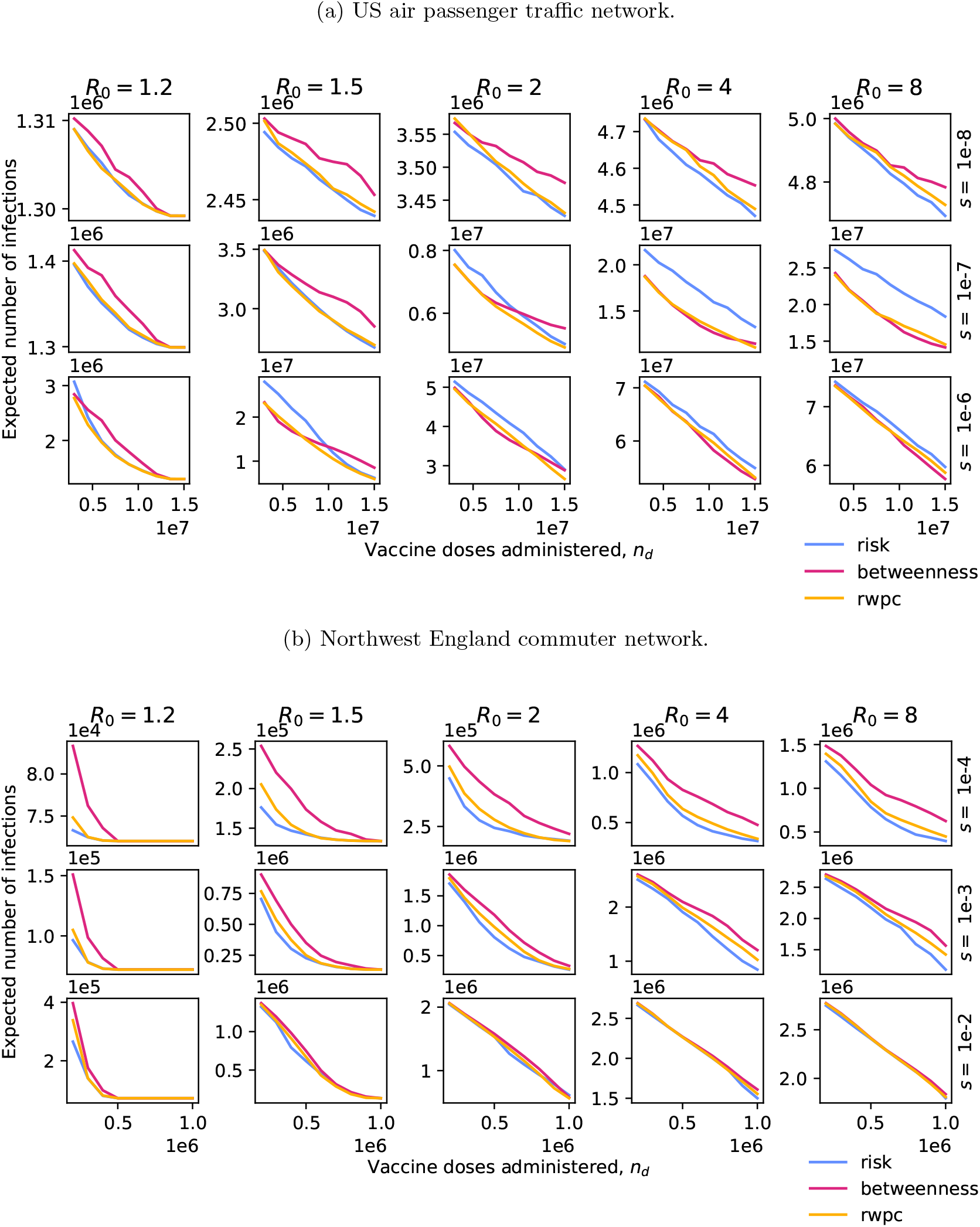
Plots of the expected total number of infections due to a disease outbreak against the number of vaccine doses administered according to one of three region prioritisation policies, for five different values of *R*_0_ and three different values of the travel scaling factor *s*, in two different networks based on empirical data.

In figure 4a, we see that the dynamics of vaccine targeting in the US air passenger traffic network mirrors that in the joined grids network, with the risk-targeting strategy performing best at low *R*_0_ and low interregional travel, and the centrality-targeting strategies performing best at high *R*_0_ and high inter-regional travel. The main differences include the overall high performance of the RWPC strategy, which is or is very close to the best strategy in all cases; the smoothness of curves, with no sharp dips in expected infections; and the convergence of the effectiveness of the various strategies for *s* = 10^*−*6^.

In figure 4b, we see a case of an empirical network in which the risk-targeting strategy is consistently the most effective at reducing infections, as seen for an artificial network in figure 3b. The effectiveness of the different strategies converges as *s* increases.

## 5 Discussion

In the Introduction, we presented a hypothesis concerning the allocation of vaccine doses to various regions— namely, that risk-targeting would perform better than centrality-targeting when *R*_0_ and inter-region travel rates were high relative to the number of vaccine doses distributed. We find that this hypothesis is borne out by our results, but with the important caveat that the network structure must allow effective use of centrality-targeting. In networks with many weak edges between clusters, the effectiveness of centrality-targeting never exceeds that of risk-targeting (see figures 3b and 4b).

This provides an important perspective on the widespread use of risk-targeting in spatial vaccination studies. Risk-targeting can perform very well in certain networks, or in certain parameter regimes, but is far from universally superior to alternative strategies. In particular, when a pathogen is spreading quickly, or when vaccine availability is limited, centrality-targeting strategies should be considered as an alternative. However, it is important to choose the correct centrality measure for the system at hand. We have shown that blunt application of a popular measure of node importance, such as betweenness, can result in an less effective allocation of vaccines compared to a measure tailored to the dynamics of a pathogen spreading on a network. In this study, we used a variation on the random walk percolation centrality measure due to Piraveenan et al. [12], but different scenarios may require the application of different measures. For instance, if the current distribution of infections is unknown, percolation centrality cannot be applied, and random walk betweenness centrality may be more effective.

The scenarios explored in this study assume a vaccination campaign that can efficiently vaccinate the selected portion of the population before a pathogen becomes widespread. The interacting dynamics of vaccination and transmission for a pathogen that is epidemic or endemic in a population differ significantly from the dynamics explored above [43, 44, 45]. Further work is required to know whether risk-targeting is the most effective strategy for combating an endemic pathogen. With SARS-CoV-2 already prevalent in most regions of the world, this means that our results are not applicable to ongoing COVID-19 vaccination efforts—however, they may provide insight into preventing the invasion of new strains of the virus, if these variants are detected early.

In our model, we assumed a single-dose vaccine with 100% efficacy and no waning immunity. A more complex model of vaccination could expand on our results in interesting ways. An expanded model could consider the different effects of vaccines—infection-reducing, disease-reducing, and/or transmission-reducing [46, 47, 48, 49, 50]. Varying efficacy in each of these dimensions could affect the results of risk-targeting and centrality-targeting strategies in different ways. Further analyses could also consider different dosing regimes (e.g. vaccines that require multiple doses for maximal efficacy) and the effects of waning vaccine- or infection-induced immunity [51, 52, 53].

Vaccination is of course only one way to prevent a pathogen from spreading, and vaccines are not always available early on in an outbreak, especially of a novel pathogen. Risk-targeting is also used when modelling the allocation of prophylactic drugs [9], or the imposition of non-pharmaceutical interventions (NPIs) [54]. In the supplementary information, we extend our framework to a preliminary model of NPIs, and show that our core results are replicated in this case. However, a more complex model is needed to effectively capture the dynamics of different NPIs, and to combine the range of NPIs and pharmaceutical interventions used during emerging outbreaks in a single model. This is a promising avenue for further work.

This study has focussed on spatial targeting, and has investigated only two types of strategy. In principle, there are as many possible vaccination strategies as there are orderings of individuals to vaccinate. We believe that risk-targeting and centrality-targeting are the best spatial strategies currently in contention, but the vast space of possible strategies is ripe for exploration. As well as variations on centrality targeting, future work could explore generalisations of our results to demographic targeting, investigating when the vaccination of at-risk individuals is most effective relative to those with high spreading capacity (e.g. those with many contacts) [55, 56, 57].

Finally, analysis of this kind could be applied to realistic models of particular infectious disease outbreaks, providing practical guidance for vaccination campaigns. These applications should consider practical restrictions to vaccine deployment, potentially including penalising vaccination strategies that have significant spatial complexity. Such work would further elucidate the circumstances in which different types of spatial vaccination targeting strategies would be effective in reducing infections during future outbreaks.

## Data Availability

All data used in this study are freely available from the sources cited in the text. The Python code written for this study is available at \url{https://osf.io/gs9cp/}.

## 6 Authors’ contributions

BJS, RNT, and MBB conceived the study. BJS carried out the analysis, wrote the manuscript, and prepared the figures. RNT and MBB supervised the research. All authors revised the manuscript and gave final approval for submission.

## 7 Funding statement

This work was supported by funding from the Biotechnology and Biological Sciences Research Council (BBSRC) [grant number BB/M011224/1].

## 8 Code availability

All data used in this study are freely available from the sources cited above. The Python code written for this study is available at https://osf.io/gs9cp/.

## Notes

### Competing Interest Statement

The authors have declared no competing interest.

### Author Declarations

University of Oxford Central University Research Ethics Committee

