## Supplementary Information for "Evaluating strategies for spatial allocation of vaccines based on risk and centrality"

September 2, 2021

#### 1 Random walk percolation centrality

In the main text we state that the random walk percolation centrality $rwp_v(t)$ of node $v$ at time $t$ is given by:

$$rwp_v(t) = \frac{1}{(n-2)} \sum_{s \neq v, r \neq v} I_v^{(s,r)} \frac{x_s(t)}{[\sum_i x_i(t)] - x_v(t)}. \tag{1}$$

This equal to the proportion of random walks, starting at $s$ and ending at $r$, pass through $v$, averaged over values of $s$ and $r$ with a weighting determined by the percolation states $x_i$ at time $t$. In what follows we will show that $I_v^{(s,r)}$ is given by the equation

$$I_v^{(s,r)} = \sum_{e: v \in e} \frac{1}{2} \left| F_{es} - F_{er} \right|, \tag{2}$$

where the sum over values of $e$ represents a sum over all edges in the network, and matrix $F$ is defined using the equation

$$F = BC^T. \tag{3}$$

$B$ is the oriented incidence matrix of the adjacency matrix $\Lambda$, whose entries $\lambda_{i,j}$ give the weight of the edge from node $i$ to node $j$. $C$ is a matrix based on the Laplacian $L$ of $\Lambda$:

$$C = \begin{pmatrix} 0 & \mathbf{0}^T \\ \mathbf{0} & \tilde{L}^{-1} \end{pmatrix}, \tag{4}$$

where, if the Lapalacian is an $n \times n$ matrix, then the *reduced Laplacian* $\tilde{L}$ is an $n-1 \times n-1$ matrix obtained from $L$ by omitting the first row and column.

We follow proofs by Ulrik Brandes and Daniel Fleischer [1], and Martin Newman [2]. Suppose a random walk in an $n$-node network with adjacency matrix $\Lambda$ is at node $v \neq r$. Then the probability $P_{vu}$ for the random walk to move to an adjacent node $u$ is:

$$p_{vu} = \frac{\lambda_{vu}}{\sum_u \lambda_{vu}}. \tag{5}$$

We denote the $n \times n$ matrix of these quantities $P$. Now, if $v = r$, then the random walk cannot escape because it has reached the sink node, so $p_{ru} = 0$ for all $u$. We write $P_r$ to denote the $(n-1) \times (n-1)$ matrix which excludes the row and column corresponding to node $r$.

The probability that a random walk starting at node $s$ reaches node $v$ after a number of steps $m$ is given by $(P_r)^m_{sv}$. The probability that the walk then steps to an adjacent node $u$ is given by $p_{vu}\,(P_r)^m_{sv}$. So the expected number of times $N_{vu}$ for the random walk to travel from node $v$ to node $u$ is given by:

$$\begin{aligned} N_{vu} &= \sum_{m=0}^{\infty} p_{vu}\left[(P_r)^m\right]_{sv} \\ &= p_{vu}\left[(\mathbb{1} - P_r)^{-1}\right]_{sv} \\ &= \lambda_{vu}\frac{1}{\sum_u \lambda_{vu}}\left[(\mathbb{1} - P_r)^{-1}\right]_{sv} \\ &= \lambda_{vu}\left[(W - \Lambda_r)^{-1}\right]_{sv}, \end{aligned} \tag{6}$$

where $\mathbb{1}$ is the $(n-1) \times (n-1)$ identity matrix, $W_{vu} = \frac{\delta_{vu}}{\sum_k \lambda_{vk}}$, and $\Lambda_r$ is $\Lambda$ with the $r^{\text{th}}$ row and column set to zero.

The net flow $I_e^{(s,r)}$ through an edge $e = \{v, u\}$ is given by the absolute difference between the flows either way along the edge,

$$I_e^{(s,r)} = \left|N_{vu} - N_{uv}\right|. \tag{7}$$

$I_e^{(s,r)}$ can be expressed in terms of $C$ when node 1 is the sink, since $L = (W - \Lambda)$ by the definition of the Laplacian. It cannot be expressed directly in terms of the Laplacian, because $L$ is singular and thus not invertible. When node 1 is the sink, $N_{vu} = \lambda_{vu}C_{sv}$. When $r \neq 1$, we can still express $I_e^{(s,r)}$ in terms of $C$ if we continue to treat node 1 as a sink for the purpose of defining $P_1$, but take away any contribution to $N_{vu}$ from paths that emanate from node $r$, thus setting node $r$ as a sink in practise. That is,

$$I_e^{(s,r)} = \left|\lambda_{vu}C_{sv} - \lambda_{vu}C_{rv} - \lambda_{uv}C_{su} + \lambda_{uv}C_{ru}\right|. \tag{8}$$

Paths that pass first through node 1 and then along edge $e$ on the way from node $s$ to node $r$ are now included as a negative term in $N_{uv}$, rather than as a positive term in $N_{vu}$. The contribution to

$I_e^{(s,r)}$ is equivalent.

The net flow $I_v^{(s,r)}$ through a node $v$ is equal to half the sum of flows along each of its connected edges, giving

$$\begin{aligned} I_v^{(s,r)} &= \sum_{e:v\in e} \frac{1}{2} \left| \lambda_{vu} C_{sv} - \lambda_{vu} C_{rv} - \lambda_{uv} C_{su} - \lambda_{uv} C_{ru} \right| \\ &= \sum_{e:v\in e} \frac{1}{2} \left| F_{es} - F_{er} \right| \end{aligned} \tag{9}$$

where $F = BC^T$, and the oriented incidence matrix $B$ is defined:

$$B_{ve} = \begin{cases} \lambda_{vu} & \text{if } e = \{v,u\} \\ -\lambda_{vu} & \text{if } e = \{u,v\} \\ 0 & \text{otherwise.} \end{cases} \tag{10}$$

Expressing $I_v^{(s,r)}$ in terms of the oriented incidence matrix and the Laplacian allows it to be calculated using standard matrix functions. To obtain equation 1, we use the same methodology as Piraveenan et al. [3], but using the current $I_v^{(s,r)}$ for a weighted, directed matrix as defined in equation 2, rather than the current for an unweighted, symmetric matrix.

### 2 The role of long-distance travel

In figure 1 we show variations on the plots in figure 3a in the main text, with the value of the long-distance travel parameter $a$ increased to 3 and 6, respectively. The broad patterns of the results are not greatly altered by the increase in $a$, but the risk-targeting strategy is overall advantaged when long-distance travel decreases.

### 3 Non-pharmaceutical interventions

We adapted the epidemiological model presented in section 2.1 to perform a preliminary exploration of spatial NPI strategies. Instead of having a fixed number of doses $n_d$ of a vaccine, we assumed that some fixed maximum number of individuals $n_r$ could be placed under transmission-reducing restrictions. We assumed that regions placed under these restrictions experience a 66% reduction in transmission rate. Unlike the case of vaccination, there is no change in the susceptibility of the population. For each strategy, we placed the entire population of the highest-priority region under restrictions, and continued down the priority ordering until placing the next region under restrictions would cause the total population under restrictions to exceed $n_r$. This last region was not placed under restrictions, meaning that the total population under restrictions could be significantly below

(a) $a = 3$

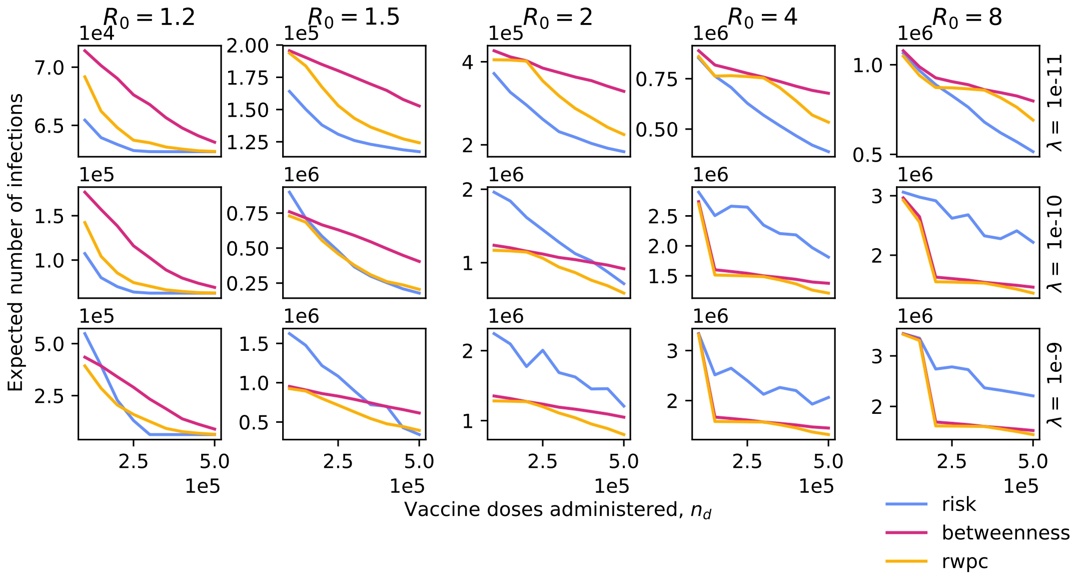


(b) $a = 6$

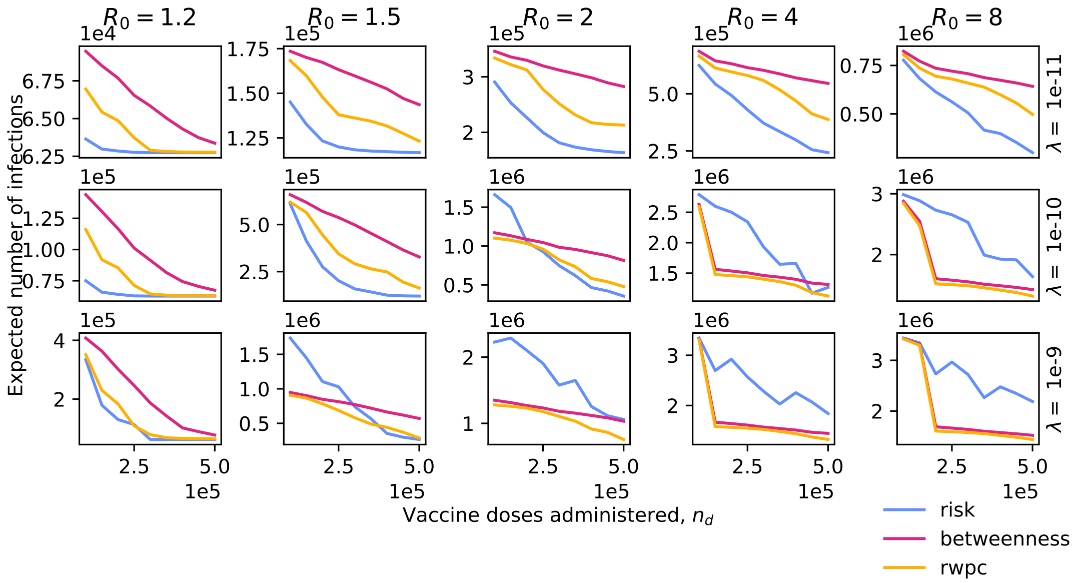


Figure 1: Plots of the expected total number of infections due to a disease outbreak against the number of vaccine doses administered according to one of three region prioritisation policies, for five different values of $R_0$ and three different overall travel rates, for two different values of the long-distance travel parameter $a$. Larger $a$ means less long-distance travel.

$n_r$, and producing the step-like shape of the plots in figure 2 as $n_r$ reaches integer multiples of the region population size.

In general, the results illustrated in figures 2 and 3 show that the relative performance of the various prioritisation strategies is similar in the case of NPIs to the case of vaccine allocation. In particular, the conclusion that risk-targeting performs better than centrality-targeting when $R_0$ and inter-region travel rates are high relative to the number of vaccine doses distributed holds out for the one-to-one joined grid network and the US air passenger traffic network, while risk-targeting almost always performs as well or better than centrality-targeting in the four corners joined grids network and the Northwest England commuter network.

(a) One-to-one joined grids network

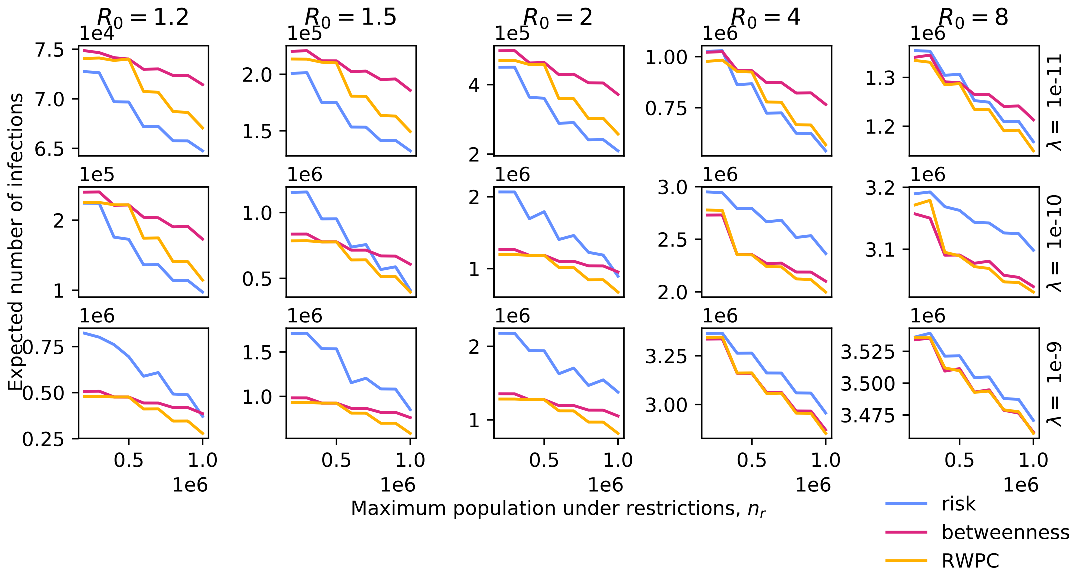


(b) Four corners joined grids network

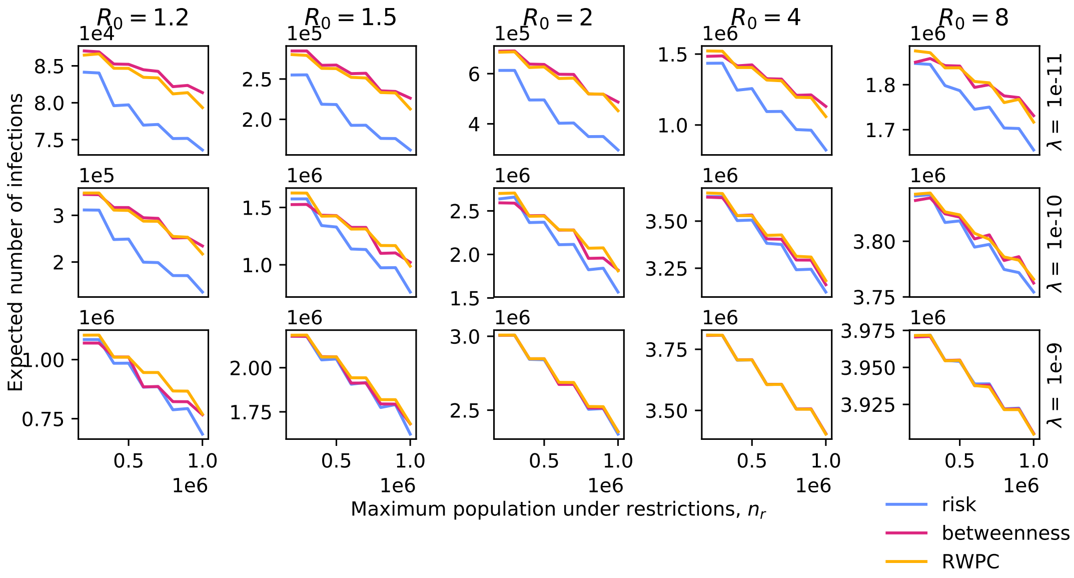


Figure 2: Plots of the expected total number of infections due to a disease outbreak against the number of individuals placed under transmission-reducing restrictions, chosen according to one of three NPI strategies, for five different values of $R_0$ and three different overall travel rates, in two variations on the 'joined grids' network.

(a) US air passenger traffic network.

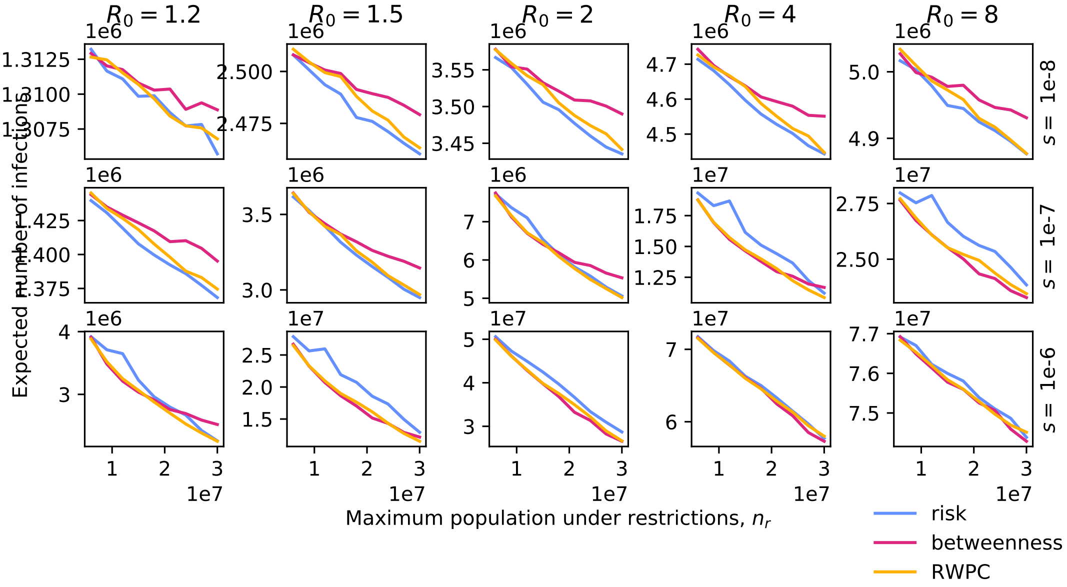


(b) Northwest England commuter network.

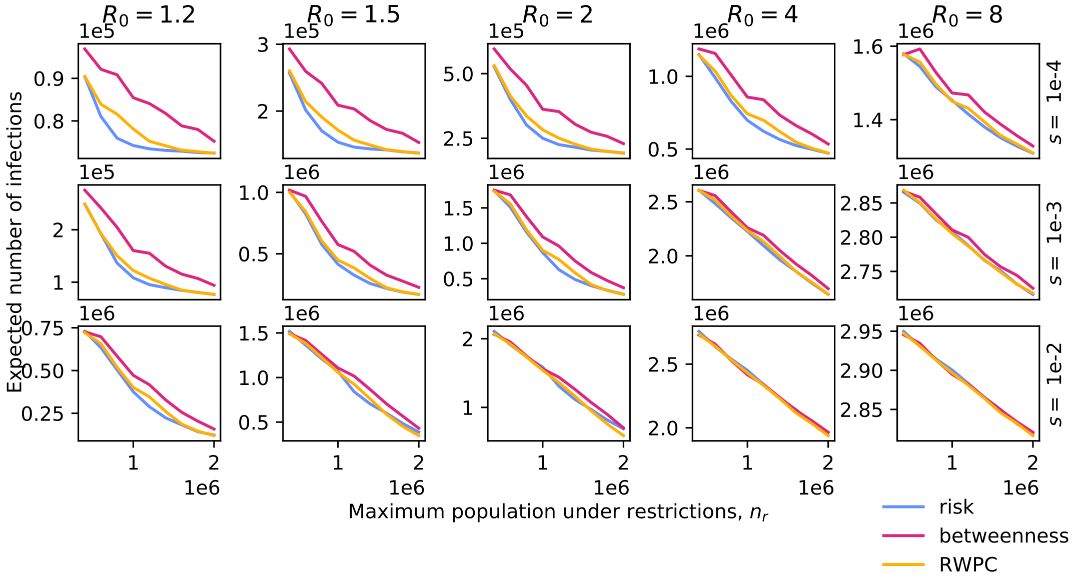


Figure 3: Plots of the expected total number of infections due to a disease outbreak against the number of individuals placed under transmission-reducing restrictions, chosen according to one of three NPI strategies, for five different values of $R_0$ and three different values of the travel scaling factor $s$, in two different networks based on empirical data.
